# Misclassification of yellow fever vaccination status revealed through hierarchical Bayesian modeling

**DOI:** 10.1101/2023.11.12.23298434

**Authors:** Quan Minh Tran, T. Alex Perkins

## Abstract

Vaccination coverage estimates are crucial inputs to decisions about investments in vaccination, yet they can be prone to inaccuracies. At the individual level, inaccuracies can be described in terms of the sensitivity and specificity of vaccination status. We estimated these quantities using a hierarchical Bayesian analysis of data from a test-negative study design with reported yellow fever vaccination status as the exposure. Our analysis accounted for the possibility of misclassification of both the exposure and the test at the country level. Across all countries, our median estimates of the sensitivity and specificity of vaccination status were 0.69 (95% credible interval [CrI]: 0.21-0.98) and 0.70 (95% CrI: 0.21-0.98), respectively. Median estimates at the country level ranged from 0.06 (95% CrI: 0.04-0.09) to 0.96 (95% CrI: 0.94-0.98) for sensitivity, and from 0.15 (95% CrI: 0.09-0.23) to 0.98 (95% CrI: 0.90-1.00) for specificity. This suggests that there is substantial misclassification of yellow fever vaccination status in general and extensive variation in misclassification across countries. Taking into account misclassification in vaccination status, we made adjustments to reported vaccination coverage and showed that reported coverage may be significantly underestimated in 10 out of 20 countries and significantly overestimated in 5 out of 20.

## Introduction

Vaccination coverage—defined as the proportion of a population that is vaccinated—is a critically important piece of information for public health. This information can be used to monitor vaccination efforts, identify areas where coverage is lower than desired, and determine potential barriers to vaccination. For example, knowing that certain age groups or regions have lower coverage can be used as a basis for prioritizing those populations for vaccination [1]. Additionally, vaccination coverage is useful for assessing the effectiveness of vaccination programs at reducing the burden of vaccine-preventable diseases [2]. It is also used as a standard to assess the strength of vaccination programs, to evaluate the accessibility of healthcare systems [3, 4], and as a criterion for financial support for vaccination programs in underresourced settings [5].

Despite its importance, there is considerable potential for error in vaccination coverage estimates. In some countries, these estimates are derived from computer-based systems [4, 6]. Though considered to be more advanced, errors in these systems can arise from mistakes in recall, recording, compiling records, or not taking into account human migration. In most low- and middle-income countries (LMICs), vaccination coverage is usually reported as an “administrative coverage estimate” [4], which is defined as the number of vaccine doses that were administered divided by the size of the target population. Errors may arise from this approach as a result of duplicated vaccination records, doses being administered but not recorded, or inaccurate estimates of the target population size [7]. Household surveys can be conducted to address these issues but may suffer from selection bias (i.e., unrepresentative households) and information bias (i.e., inaccurate data transcription or recall) [4, 8, 9].

One disease for which vaccination coverage estimates are particularly important is yellow fever. Although most contemporary cases are thought to result from zoonotic spillover [10], yellow fever is capable of large outbreaks with high mortality [11, 12]. As a result, the World Health Organization (WHO) has dedicated an initiative to the elimination of yellow fever epidemics [13]. Because there is a highly efficacious vaccine for this disease and few other effective tools for prevention [14], achieving high vaccination coverage in at-risk populations is essential. Understanding of vaccination coverage for yellow fever may be impaired by the fact that it poses the greatest risk to rural populations in Africa and South America, where accurate estimation of vaccination coverage is challenging [7, 15]. This emphasizes the need for accurate estimates of yellow fever vaccination coverage.

In this study, we estimated the sensitivity and specificity of vaccination status using a test-negative, case-control study design involving surveillance data on yellow fever in Africa. Similar approaches have been taken previously to estimate vaccine effectiveness in light of diagnostic inaccuracy [16, 17, 18, 19], exposure inaccuracy [20], or both [21]. Our analysis allowed for the possibility of inaccuracy in both the exposure (vaccination status) and the test (infection status), not to adjust estimates of vaccine effectiveness but to obtain estimates of vaccination misclassification in a real-world setting. Additionally, we assessed the extent of geographic variability in misclassification by comparing alternative models with differing assumptions about geographic variability in the sensitivity and specificity of vaccination status, the sensitivity and specificity of diagnostic testing, the proportion of infections that were reported, and the force of infection. Finally, we showed how estimates of sensitivity and specificity of vaccination status can be used to adjust vaccination coverage estimates to account for vaccination status misclassification.

## Methods

### Data

We used a database of yellow fever surveillance records from Africa collated by the World Health Organization. This database consists of suspected cases of yellow fever (based primarily on a case definition of fever and jaundice [22]), of which there were approximately 15,000 in the database from 20 African countries between 2005 and 2011. Suspected cases who tested positive for yellow fever virus immunoglobulin M were classified as confirmed cases of yellow fever. Each individual’s age and reported history of vaccination against yellow fever were also recorded. We used spatial information about each case at the first administrative level (adm1). Use of this database for this analysis was deemed not human subjects research by the University of Notre Dame Institutional Review Board (Protocol 22-04-7193). For our main analysis, we used yellow fever vaccination coverage estimates by age and year [23]. These estimates span 1950 to 2050 and provide information on vaccination coverage at the adm1 level.

### Model

Each suspected case of yellow fever in the database was classified according to two observed states: *C* (1 = test-positive, 0 = test-negative) and *R* (1 = reported vaccinated, 0 = reported non-vaccinated). Our model also considered two latent states for each suspected case: *I* (1 = truly infected, 0 = truly not infected) and *T* (1 = truly vaccinated, 0 = truly not vaccinated). These states are related in our model according to causal dependencies that describe how the data were generated (Fig. 1).

**Figure 1.**
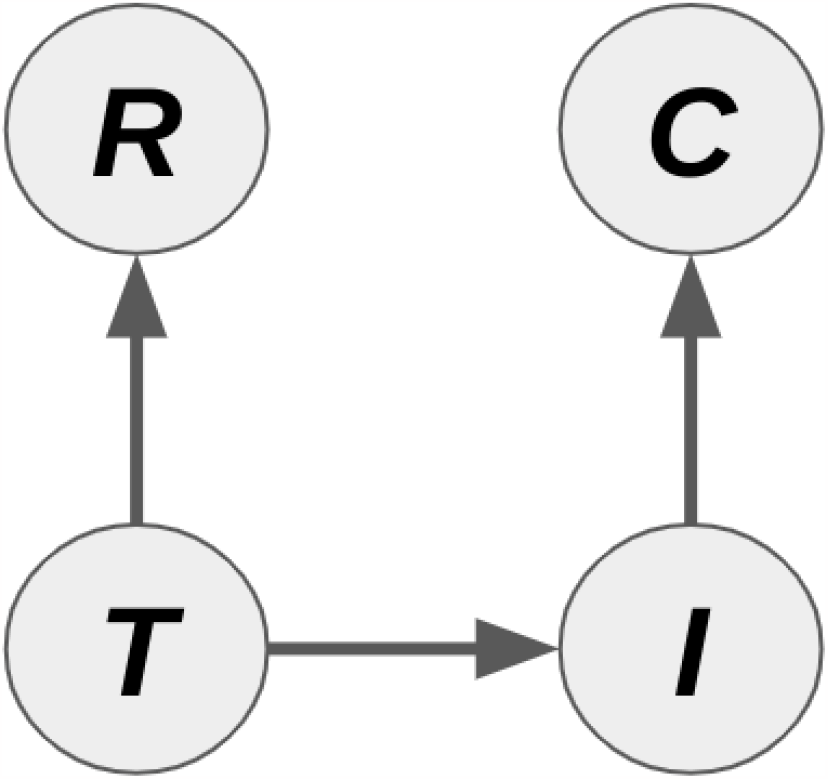
Vaccination status misclassification model. The diagram shows the causal relationships among four states for each individual in the database: *R*, reported vaccination status; *T*, true vaccination status; *I*, true infection status; and *C*, reported disease status. The core of the model’s likelihood, *ℒ*_*data*_, depends on probabilities of each of these three causal relationships. Pr(*R*|*T*) depends on parameters describing the sensitivity (*Se*_*vac*_) and specificity (*Sp*_*vac*_) of vaccination classification, Pr(*I*|*T*) depends on the force of infection (*λ*) and reporting probability (*ρ*), and Pr(*C*|*I*) depends on parameters describing the sensitivity (*Se*_*test*_) and specificity (*Sp*_*test*_) of case confirmation.

First, *T → R* is determined by the sensitivity and specificity of reported vaccination status in each country *k*, which we represent with *Se*_*vac,k*_ and *Sp*_*vac,k*_, respectively. Specifically,

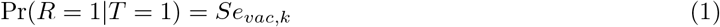

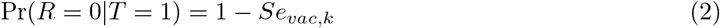

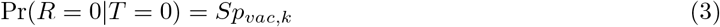

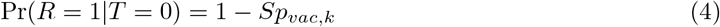

describe the relationships among all combinations of *T* and *R*. We enforced a constraint on *Se*_*vac,k*_ and *Sp*_*vac,k*_ to ensure that *Se*_*vac,k*_ +*Sp*_*vac,k*_ *>* 1, because the alternative would imply that vaccination status is, on average, misclassified more often than not. To implement this constraint, we modeled 1 − *Se*_*vac,k*_, 1 − *Sp*_*vac,k*_, and *Se*_*vac,k*_ + *Sp*_*vac,k*_ − 1 as random effects across countries *k* following a Dirichlet distribution. In other words, we assumed that the country-specific values of these parameters follow a global Dirichlet distribution with concentration parameters *α*_*vac,1*_, *α*_*vac,2*_, and *α*_*vac,3*_.

Second, *T* → *I* is determined by the probability of contracting yellow fever given one’s true vaccination status, which we represent with Pr(*I*|*T*). We start with the case where *T* = 0 and *I* = 1, the probability of which is

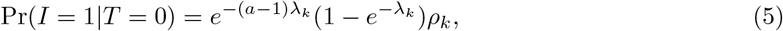

where *a* is age at the time that an individual is recorded as a suspected case, *λ*_*k*_ is the force of infection of yellow fever virus in the individual’s country *k*, and *ρ*_*k*_ is the probability that an infection is reported in country *k*. The term 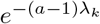 represents the probability that an individual avoids infection before age *a*, whereas 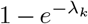 represents the probability that the individual becomes infected at age *a*. This formulation is widely known within epidemiology as a “catalytic” model [24] and has been used in prior models of yellow fever disease burden [22, 25, 26]. Eqn. (5) implies that

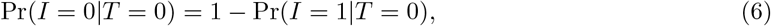

which encompasses all the ways in which an individual would not become a suspected case with an etiology of yellow fever virus at age *a*. Individuals who are truly vaccinated become suspected cases with an etiology of yellow fever virus with probability

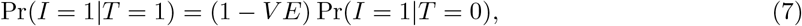

where *V E* is vaccine efficacy. Similar to eqn. (6),

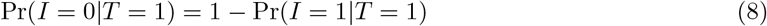

is the probability that a truly vaccinated person does not become a suspected case with an etiology of yellow fever virus at age *a*.

Third, *I → C* is determined by the sensitivity and specificity of laboratory testing in each country *k*, which we represent with *Se*_*test,k*_ and *Sp*_*test,k*_, respectively. Like eqns. (1)-(4),

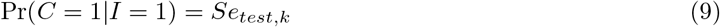

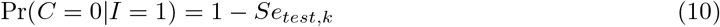

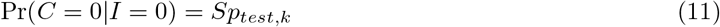

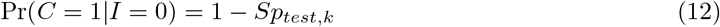

describe the probabilities that confirmed cases result from suspected cases with differing true infection statuses in country *k*. We note that individuals with a status of *I* = 1 represent reported infections only, given that the reporting probability *ρ*_*k*_ filters out infections that went unreported in country *k*. We treat *Se*_*test,k*_ and *Sp*_*test,k*_ as fixed effects.

Based on these three causal dependencies, we can define the joint distribution of all four states (*c, i, r, t*) for each individual to be

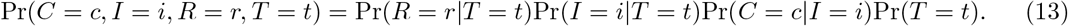

All of the terms on the right-hand side of eqn. (13) have already been defined except for Pr(*T* = *t*), which pertains to the latent state of true vaccination status. To define this, we begin by specifying the relationship between Pr(*T* = 1) and Pr(*R* = 1), which is

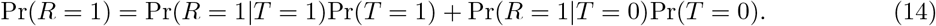

Solving for Pr(*T* = 1), we obtain

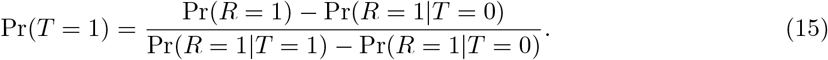

As Pr(*R* = 1|*T* = 1) and Pr(*R* = 1|*T* = 0) have already been defined, that leaves only Pr(*R* = 1) to be defined. For this quantity, we used vaccination coverage estimates appropriate to each suspected case’s home adm1, year, and age. Finally, to obtain a probability of the observed states only, we marginalize over states *T* and *I* in eqn. (13) to obtain

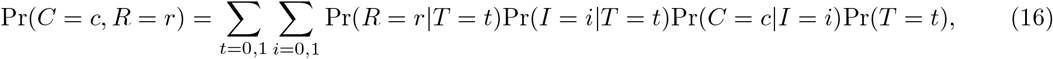

which is the joint probability of the observed states (*c, r*) for each individual.

The likelihood of the model and its parameters consists of two parts. First, the contribution to the likelihood from data on the confirmed case (*c*_*j*_) and reported vaccination (*r*_*j*_) status across all individuals *j* is

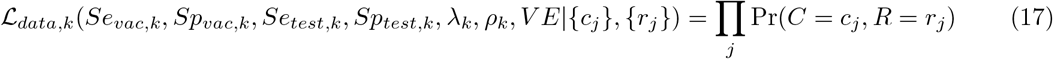

for country *k*. The variables {*c*_*j*_} and {*r*_*j*_} refer to the status of *c* and *r* across the set of individuals *j* within country *k*. The data in this analysis ultimately consist of counts of the number of individuals within each country with each possible combination of values of *c* (0 or 1) and *r* (0 or 1), because that determines how many times each Pr(*C* = *c, R* = *r*) from the four possible combinations of *c* and *r* is multiplied in the likelihood in eqn. (17). Our analysis constitutes a test-negative design in the sense that it utilizes information on the proportions of individuals with a given vaccination status (*r*) in groups distinguished by their case confirmation status (*c*). Differences in the proportions between these groups provide information about the parameters, with *Se*_*vac*_ and *Sp*_*vac*_ being of primary interest.

Second, the contribution to the likelihood from the distribution of country-specific sensitivities and specificities of reported vaccination status is

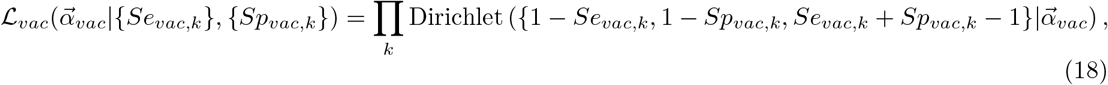

where 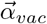 contains three concentration parameters, *α*_*vac,1*_, *α*_*vac,2*_, and *α*_*vac,3*_. This component of the likelihood allows for a balance to be achieved between the freedom of the *Se*_*vac,k*_ and *Sp*_*vac,k*_ parameters to vary among countries and the constraint that similar combinations of those parameters may apply to multiple countries. Mathematically, this balance is achieved by allowing the country-specific parameter values to follow a common distribution. The choice of a Dirichlet distribution was motivated by a desire to model the *Se*_*vac,k*_ and *Sp*_*vac,k*_ parameters jointly and to ensure that they were restricted to values between 0 and 1. The third implied parameter of this Dirichlet distribution, *Se* + *Sp* − 1, allowed us to specify a prior for the overall accuracy of case confirmation.

Together, these component likelihoods lead to

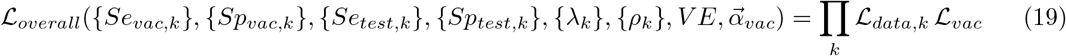

as the overall likelihood of the model and its parameters. Notation such as *{Se*_*vac,k*_*}* refers to the collection of *Se*_*vac,k*_ parameters across the set of countries *k*, and similar for other parameters specified for each country. In contrast to *Se*_*vac,k*_ and *Sp*_*vac,k*_, the country-specific parameters *Se*_*test,k*_, *Sp*_*test,k*_, *λ*_*k*_, and *ρ*_*k*_ were not modeled as belonging to common distributions and were allowed to vary freely among countries. Posterior estimates of these parameters were found to closely follow their prior distributions, suggesting that modeling them as belonging to a common distribution would have had only a negligible impact on our results.

### Priors

We specified informative priors for each parameter to the extent possible based on independently available information. For *λ*_*k*_, we used posterior estimates of force of infection at the country level from a previous study [26]. Because that study produced separate posterior estimates under eight alternative scenarios, we fitted a normal distribution on the log_*10*_ scale to posterior samples of force of infection for each country pooled across the eight scenarios. For *ρ*_*k*_, we used posterior estimates from the same study [26] to inform our priors, albeit with the same prior for each country *k* since that study only obtained a single estimate of reporting probability. In this case, we fitted a normal distribution to logit-transformed values of reporting proportion estimates pooled across the eight scenarios. For *V E*, we used a posterior estimate from a previous study [27] to inform a beta-distributed prior, the parameters of which were obtained by matching to the previous study’s median estimate of 0.975 and 95% credible interval of 0.83 - 1.0. For *Se*_*vac*_ and *Sp*_*vac*_, we restricted the intensity parameters *α*_*vac*_ to be larger than 1 to ensure a unimodal shape to the inferred distribution of *Se*_*vac*_ and *Sp*_*vac*_ across countries. Specifically, the priors of these parameters were all exponential(1) + 1. For *Se*_*test,k*_ and *Sp*_*test,k*_, we selected values of Dirichlet intensity parameters that resulted in a median of 0.95 and 95% intervals of the marginals of *Se*_*test,k*_ and *Sp*_*test,k*_ equal to 0.8 - 1.0 and 0.6 - 1.0, respectively [28, 29, 30]. This was obtained with Dirichlet intensity parameters of 0.145, 0.569, and 5.359 for 1 − *Se*_*test,k*_, 1 − *Sp*_*test,k*_, and *Se*_*test,k*_ + *Sp*_*test,k*_ − 1.

### Hamiltonian Monte Carlo

We used the likelihood from eqn. (19) and the priors described above as the basis for obtaining Bayesian estimates of the model’s parameters using Hamiltonian Monte Carlo implemented through the rstan package [31] in R 4.2.1 [32]. We ran the models with 10^*5*^ iterations and a burn-in period of 5 *×* 10^*4*^ iterations. Convergence was assessed using 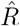 estimates from the rstan package [31] and trace plots for all parameters. We specified the threshold for convergence to be 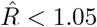.

### Model comparison

To account for uncertainty in some of the choices we made in the design of our model, we compared our full model described above to 15 other models that represent simpler cases of it. Specifically, we constructed alternative models that simplified the parameters that we allowed to vary geographically, with the alternative models treating different combinations of geographically variable parameters as constants. For example, instead of considering country-specific values of *Se*_*vac,k*_ and *Sp*_*vac,k*_, we considered an alternative with the same *Se*_*vac*_ and *Sp*_*vac*_ applying to all countries. We did the same for *Se*_*test*_ and *Sp*_*test*_, *λ*, and *ρ*. This resulted in a total of 16 different models that explored all combinations of the ways in which geographic variability could be toggled on or off across these four sets of parameters. We compared these 16 models on the basis of the Watanabe-Akaike information criterion (WAIC). We were also interested in assessing how parameter estimates differed across models.

## Results

### Model comparison

We fitted all 16 models to the data and determined that all models converged, as evidenced by 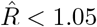 and visual inspection of trace plots. We found that the default model, which is the most complex model with four sets of geographically variable parameters, performed among the best on the basis of WAIC (Fig. 2). We observed minimal differences in WAIC among models that did or did not allow for geographic variability in force of infection or reporting proportion. Notably, geographic variability in *Se*_*vac*_ and *Sp*_*vac*_ was the most important model feature for enabling a low WAIC. This was followed by geographic variability in *Se*_*test*_ and *Sp*_*test*_ as the second most important model feature, but to a lesser extent (Fig. 2).

**Figure 2.**
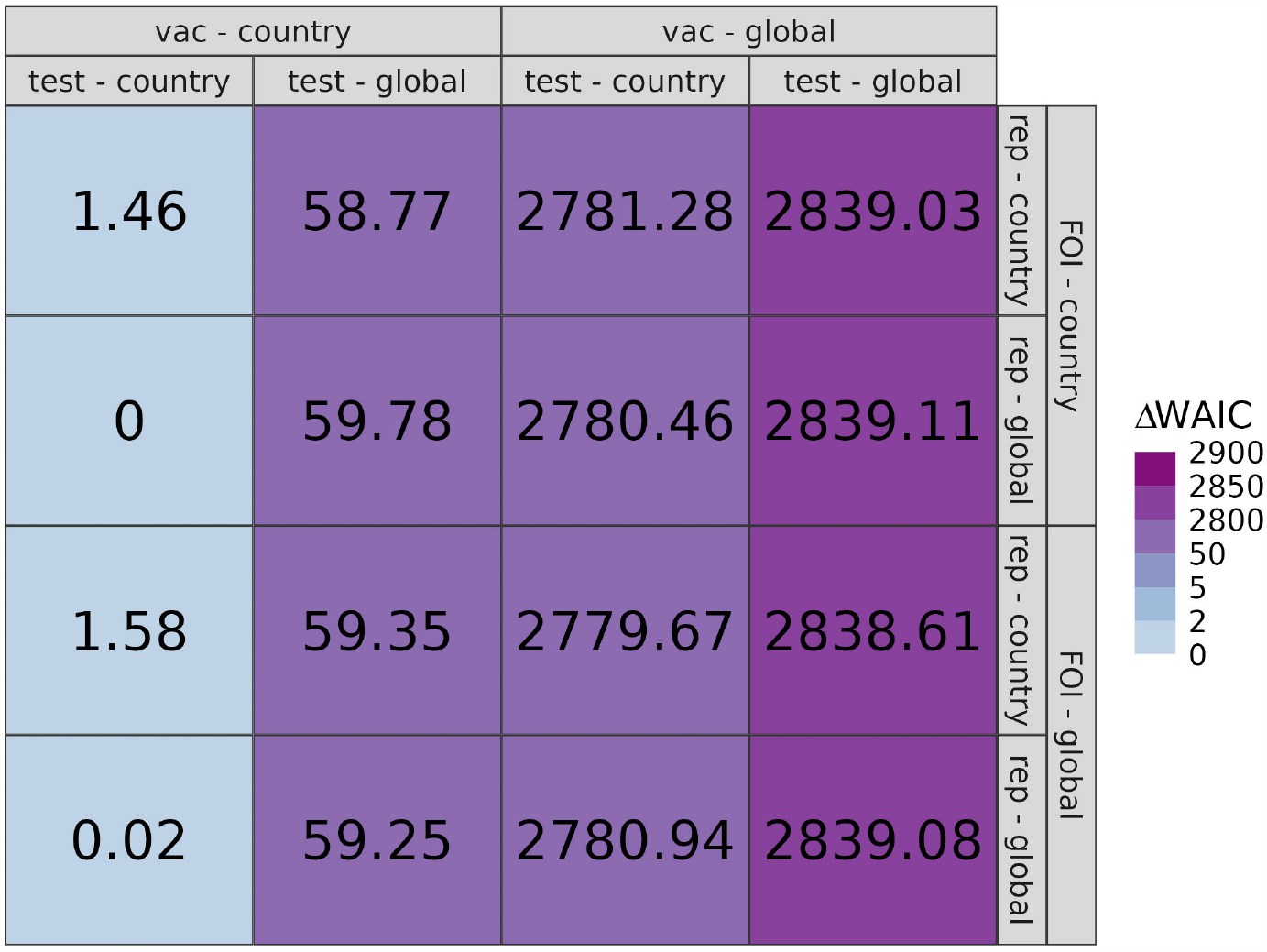
ΔWAIC values of the 16 models. ΔWAIC is defined as the difference between the WAIC value of a given model and that of the model with the lowest WAIC value. The default model (top left) includes geographic variability in *Se*_*vac*_ and *Sp*_*vac*_ (vac), *Se*_*test*_ and *Sp*_*test*_ (test), force of infection (FOI), and reporting proportion (rep). In the columns and rows, geographic variability is indicated by whether the parameters vary by country or do not vary and are thus global constants.

### Estimation of *Se*_*vac*_ and *Sp*_*vac*_

The default model showed considerable variability in estimates of the sensitivity and specificity of vaccination status across countries, with median estimates of 0.69 (95% CrI: 0.21 - 0.98) and 0.70 (95% CrI: 0.21 - 0.98), respectively. Across the 20 countries that we considered, country-specific estimates ranged from medians of 0.15 (95% CrI: 0.09 - 0.23) to 0.98 (95% CrI: 0.90 - 1.0) for sensitivity, and from medians of 0.06 (95% CrI: 0.04 - 0.09) to 0.96 (95% CrI: 0.94 - 0.98) for specificity (Fig. 3). The Pearson correlation between *Se*_*vac,k*_ and *Sp*_*vac,k*_ across countries had a median value of –0.85 (95% CrI: –0.924 - –0.711), indicative of a trade-off between the sensitivity and specificity of differing practices of vaccination status classification among countries. Notably, estimates of *Se*_*vac,k*_ and *Sp*_*vac,k*_ for several countries fell near the diagonal line, suggesting that classification of vaccination status was only marginally better than chance.

**Figure 3.**
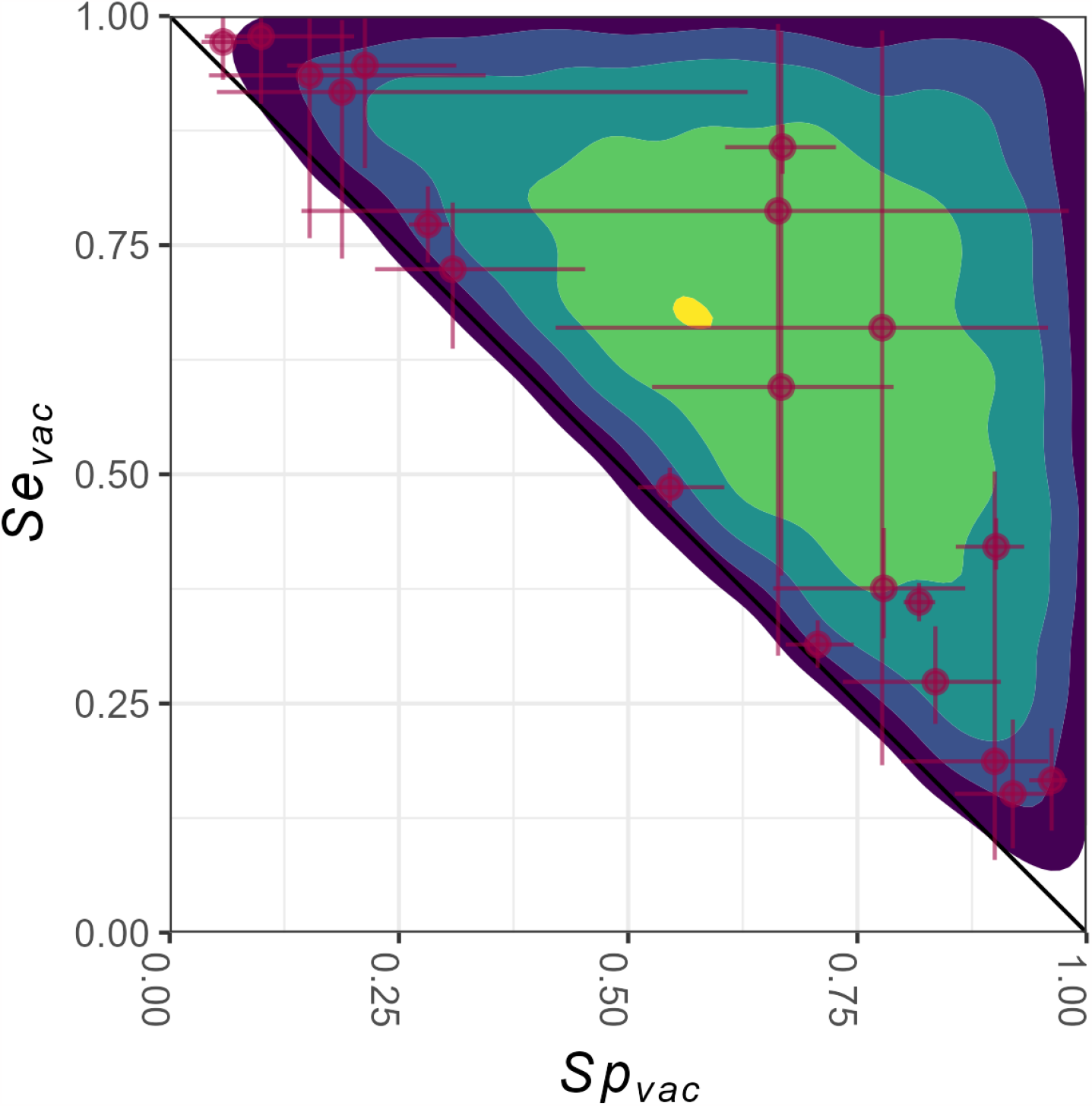
Estimates of *Se*_*vac*_ and *Sp*_*vac*_ from the default model. The points with their lines represent the median estimates and the 95% CrI of country-specific estimates *Se*_*vac,k*_ and *Sp*_*vac,k*_. The shades indicate the density of *Se*_*vac*_ and *Sp*_*vac*_ at a global level, where lighter shades indicate higher densities.

We observed that the amount of uncertainty associated with estimates of country-specific values of *Se*_*vac,k*_ and *Sp*_*vac,k*_ was influenced by the number of data points from each country. Specifically, countries with very few samples (i.e., ≤ 6) exhibited markedly higher levels of uncertainty in their estimates (Fig. S1). To assess the robustness of our results to influences from particular countries, we performed a sensitivity analysis in which we excluded the top one to five countries with the most data points. The results of this analysis indicated that overall estimates of the distribution of *Se*_*vac*_ and *Sp*_*vac*_ across countries remained largely unchanged (Fig. S2), supporting the robustness of our primary results.

Among the eight alternative models that allowed for geographic variability in *Se*_*vac,k*_ and *Sp*_*vac,k*_, we found minimal differences in estimates of *Se*_*vac,k*_ and *Sp*_*vac,k*_ (Fig. S3). The negative correlation between *Se*_*vac,k*_ and *Sp*_*vac,k*_ across countries that we observed for the default model was also apparent for the other seven models (Table S1, Fig. S4). For models that lacked geographic variability in *Se*_*vac*_ and *Sp*_*vac*_, estimates of *Se*_*vac*_ and *Sp*_*vac*_ were all similar. For example, the simplest model (Fig. S3, bottom right), yielded median estimates of *Se*_*vac*_ and *Sp*_*vac*_ of 0.49 (95% CrI: 0.48 - 0.50) and 0.71 (95% CrI: 0.70 - 0.72), respectively.

To validate our inferences, we repeated our inference procedure with the default model on ten data sets simulated with parameters set to the medians of their marginal posterior distributions. In doing so, we observed broad consistency between the country-level estimates of *Se*_*vac,k*_ and *Sp*_*vac,k*_ and their simulated values (Fig. S5). In addition to alignment in the central tendencies of the posteriors obtained from simulated and empirical data sets, the spans of their uncertainty intervals were also similar.

### Estimation of other parameters

Unlike *Se*_*vac,k*_ and *Sp*_*vac,k*_, estimates of *Se*_*test,k*_ and *Sp*_*test,k*_ varied negligibly across countries in the default model (Fig. S6). Specifically, the country-specific *Se*_*test,k*_ estimates ranged from medians of 0.99 (95% CrI: 0.82 - 1.00) to 1.00 (95% CrI: 0.81 - 1.00), and *Sp*_*test,k*_ estimates ranged from medians of 0.96 (95% CrI: 0.66 - 1.00) to 1.00 (95% CrI: 0.99 - 1.00). We consistently observed a similar pattern across all seven country-specific models, where the estimates for each country were similar across different models (Fig. S7, eight left panels). For the simplest model (Fig. S7, bottom right), which assumes no geographic variation in any parameter, the median estimates of *Se*_*test*_ and *Sp*_*test*_ were 0.99 (95% CrI: 0.82 - 1.00) and 0.99 (95% CrI: 0.98 - 0.99), respectively. In general, we observed that all estimates of *Se*_*test*_ and *Sp*_*test*_ overlapped significantly with their prior distributions, which had medians of 0.98 (95% CrI: 0.77 - 1.00) and 0.97 (95% CrI: 0.58 - 1.00), respectively. Posterior estimates of other parameters—i.e., *ρ*_*k*_, *λ*_*k*_, and *V E*—also closely resembled their prior distributions (Figs. S8-S10).

Given that our model allows for multiple sources of error in observed data, there is potential for some parameter combinations to not be identifiable. To assess the ability of the model to obtain separate, uncorrelated estimates of the sensitivities and specificities of vaccination status and testing, we first examined the correlations of their posterior samples within each of the 20 countries in our analysis. Correlations between *Se*_*vac,k*_ and *Se*_*test,k*_ ranged from −0.10 to 0.09, as did correlations between *Sp*_*vac,k*_ and *Sp*_*test,k*_. As a further test of the identifiability of these parameters, we generated another ten simulated data sets, but with lower values of *Se*_*test,k*_ and *Sp*_*test,k*_ (0.70 for both) than their median posterior estimates. Even under these lower values of *Se*_*test,k*_ and *Sp*_*test,k*_, our inferences of *Se*_*vac,k*_ and *Se*_*test,k*_ remained similar (Fig. S11).

We also explored the extent to which misestimation of other parameters—namely, the force of infection, *λ*_*k*_—could bias estimates of *Se*_*vac,k*_ and *Sp*_*vac,k*_. To test this, we simulated ten data sets with values of *λ*_*k*_ varying randomly from 10^−*5*^ to 10^−*1*^ by factors of 10 across the 20 countries in our analysis. When we applied our inference method to those data, we found that estimates of *Se*_*vac,k*_ and *Sp*_*vac,k*_ were similar to those from our primary analysis (Fig. S11). Moreover, estimates of *λ*_*k*_ also resembled those from the primary analysis (Fig. S12). In both sets of analyses, posterior estimates of *λ*_*k*_ were driven strongly by their priors.

### Vaccination coverage adjusted for misclassification

By using the default model estimates of *Se*_*vac,k*_ and *Sp*_*vac,k*_, we were also able to adjust for bias in estimates of vaccination coverage. We did this by calculating adjusted vaccination coverage using eqn. (15), where reported vaccination coverage comes from population-weighted vaccination coverage estimates from POLICI between 1980 and 2023 (see Fig. 4).

**Figure 4.**
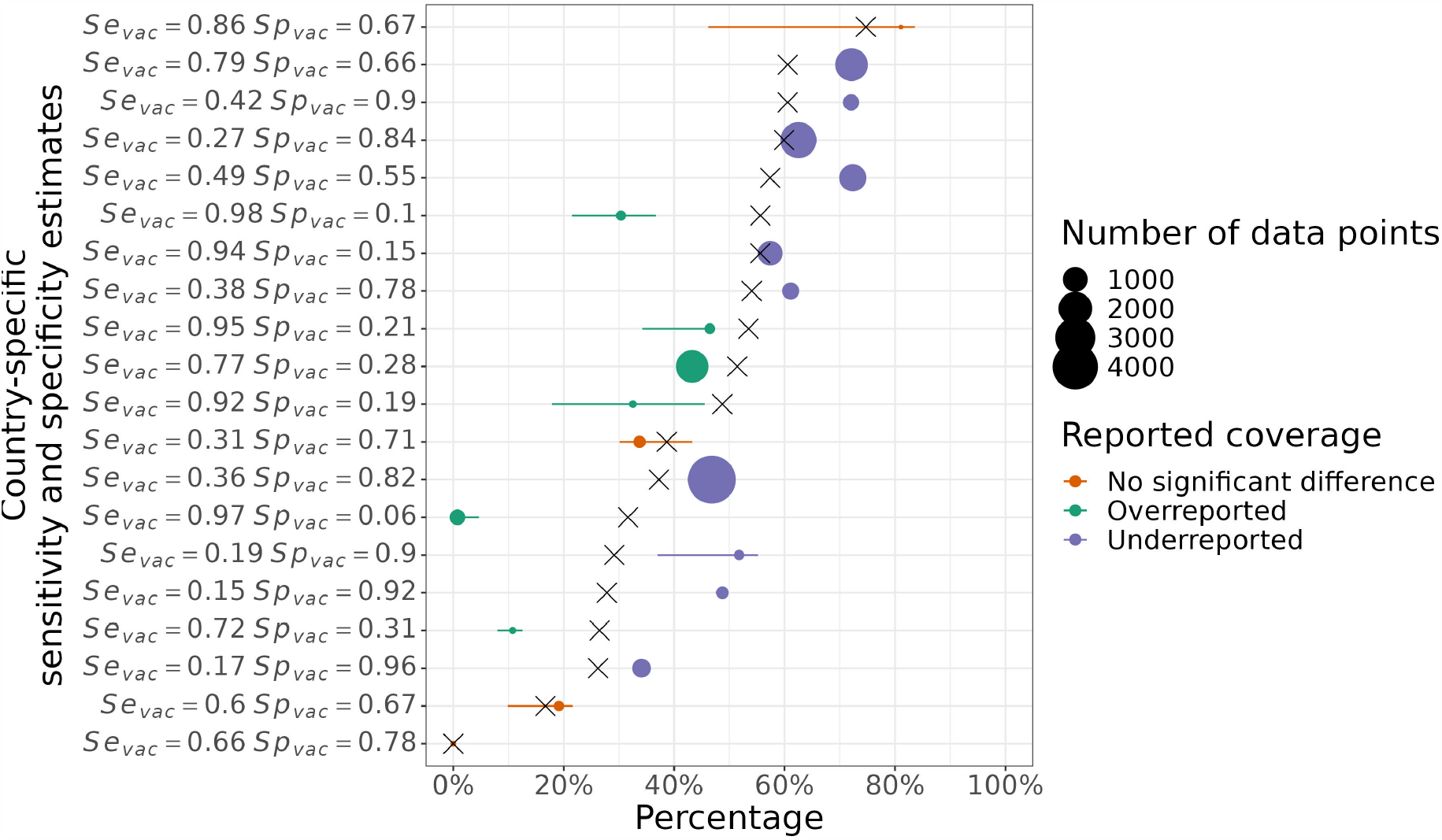
Reported versus adjusted vaccination coverage. Reported vaccination coverages from POLICI are shown with an “X”. The horizontal points and lines indicate the medians and 95% posterior predictive intervals of adjusted vaccination coverage, colored by the direction of bias (underestimate, overestimate, or no significant difference). The size of each point corresponds to the number of data points from each country. The countries are arranged from bottom to top based on their reported vaccination coverage. The sensitivity and specificity of vaccination status for each country is shown on the left.

Our findings revealed that adjusted vaccination coverage could either increase or decrease relative to reported coverage, depending on country-specific estimates of *Se*_*vac,k*_ and *Sp*_*vac,k*_ (Fig. 4). Specifically, our analysis revealed that in ten countries, the 2.5% quantile of adjusted vaccination coverage exceeded reported coverage, implying underestimation of reported vaccination coverage. These countries exhibited a pattern of low *Se*_*vac,k*_ and high *Sp*_*vac,k*_. In five countries, the 97.5% quantile of adjusted vaccination coverage was lower than reported coverage, implying overestimation of reported vaccination coverage. These countries exhibited a pattern of high *Se*_*vac,k*_ but low *Sp*_*vac,k*_. In some countries, we noted that bias of reported vaccination coverage may be negligible, such as when reported coverage is very low. Uncertainty around adjusted vaccination coverage was driven by the degree of uncertainty about *Se*_*vac,k*_ and *Sp*_*vac,k*_, which was in turn influenced by the number of data points from each country (Fig. 4).

## Discussion

Our study aimed to quantify the degree of misclassification associated with yellow fever vaccination status using a test-negative case-control study design with surveillance data on yellow fever in Africa. We found that vaccination status misclassification appears to be prevalent, with country-level estimates of sensitivity and specificity of vaccination status being highly negatively correlated and with the average of sensitivity and specificity generally not exceeding 0.70. By quantifying these inaccuracies, we were able to obtain adjusted estimates of vaccination coverage that account for vaccination status misclassification. Our estimates of adjusted vaccination coverage indicated that reported vaccination coverage may be an underestimate in ten out of twenty countries, an overestimate in five, and approximately correct in five.

We observed considerable variability in our estimates of the sensitivity and specificity of vaccination status across the 20 countries that we considered, which could be attributable to differences in vaccination recording systems across those countries. To remedy the unique issues with each country’s vaccination recording system and improve the quality of vaccination coverage data, the WHO has developed guidelines to help identify weaknesses in vaccination recording systems and improve them [33, 34, 35]. Whatever the specific causes of this country-level variation are, this variation is unlikely to be explainable based on any readily available or predictable factors. As a result, it would be difficult to extrapolate our estimates to countries not included in our analysis, and efforts should be made to extend our analysis with additional data.

Analyses of simulated data suggest that inferences of the sensitivity and specificity of vaccination status are valid but that inferences of other parameters closely mirror prior assumptions. A primary driver of this asymmetry in parameter identifiability is that our data—which ultimately come down to the proportion vaccinated among test-negative versus test-positive individuals—are influenced more by the sensitivity and specificity of vaccination status than other parameters. Given that the data are limited to suspected cases who underwent confirmatory testing, they simply do not contain much information about other parameters, such as the force of infection or reporting probability. Reassuringly, the prior distributions for those parameters were well-informed by a previous study [26], for which estimation of those parameters was of primary interest.

Using our estimates of the sensitivity and specificity of vaccination status, we quantified discrepancies between reported and adjusted vaccination coverage. Estimates of vaccination coverage are important for estimating the burden of yellow fever, because of their implications for the use of serological data to infer the force of infection [22]. Because seropositivity as a result of natural infection or vaccination cannot be differentiated empirically, estimates of vaccination coverage are important for determining how much of measured seropositivity is attributable to natural infection [26]. If vaccination coverage is higher than reported, then force of infection may be overestimated. Conversely, if vaccination coverage is lower than reported, then force of infection may be underestimated. Such biases in estimates of the force of infection are expected to lead to corresponding biases in estimates of disease burden, which could be consequential for prioritizing investments in different vaccine-preventable diseases [2].

There are important limitations of our analysis to note, especially in relation to the state of currently available data. Of the more than 15,000 suspected cases in the database that we analyzed, only 206 were confirmed yellow fever cases. Within the same timeframe of 2008 to 2013 in the 20 countries included in our database, a total of 3,590 confirmed cases were reported by another source [36], suggesting that the data we analyzed may be very incomplete. In addition, there is substantial imbalance in the sample sizes from different countries, which could affect our global estimates of the sensitivity and specificity of vaccination status. At the same time, our sensitivity analysis suggested that omission of data from countries with the most data resulted in minimal changes to our global estimates. There was, nevertheless, high uncertainty in parameters for countries with low sample sizes. Moreover, the database that we used includes only 20 of the more than 30 countries in Africa where there is a risk of yellow fever. There are also limitations of the vaccination coverage estimates that we used to inform Pr(*R* = 1). Reported vaccination coverage may vary at finer temporal and spatial scales than the estimates available to us, meaning that the values of Pr(*R* = 1) that we used may be imperfect approximations and could potentially lead to bias in our estimates of *Se*_*vac,k*_ and *Sp*_*vac,k*_.

In the future, the analytical framework we used could be applied to evaluate country-level vaccination reporting systems for other diseases. The requirements for doing so would be the same type of data on vaccination and infection statuses for suspected cases of whatever disease is of interest.

Replication of our analysis for other diseases could shed light on the consistency (or not) of the sensitivity and specificity of vaccination status across multiple diseases, leading to a unique approach for comprehensive assessment of a given country’s vaccination recording system. Accurate understanding of vaccination coverage is important for achieving public health goals for a variety of other infectious diseases, such as measles [37] and polio [38].

## Data Availability

Data are not available due to the fact that they were made available to the study authors by a third party through a data-sharing agreement with the data provider that does not allow data sharing by the authors.

## Acknowledgements

We acknowledge the World Health Organization for sharing its yellow fever surveillance database for use in this analysis. This work was carried out as part of the Vaccine Impact Modelling Consortium (www.vaccineimpact.org), but the views expressed are those of the authors and not necessarily those of the Consortium or its funders. The funders were given the opportunity to review this paper prior to publication, but the final decision on the content of the publication was taken by the authors. This work was supported, in whole or in part, by the Bill & Melinda Gates Foundation, via the Vaccine Impact Modelling Consortium [Grant Number INV-009125]. Under the grant conditions of the Foundation, a Creative Commons Attribution 4.0 Generic License has already been assigned to the Author Accepted Manuscript version that might arise from this submission.

---

**Figure S1:**
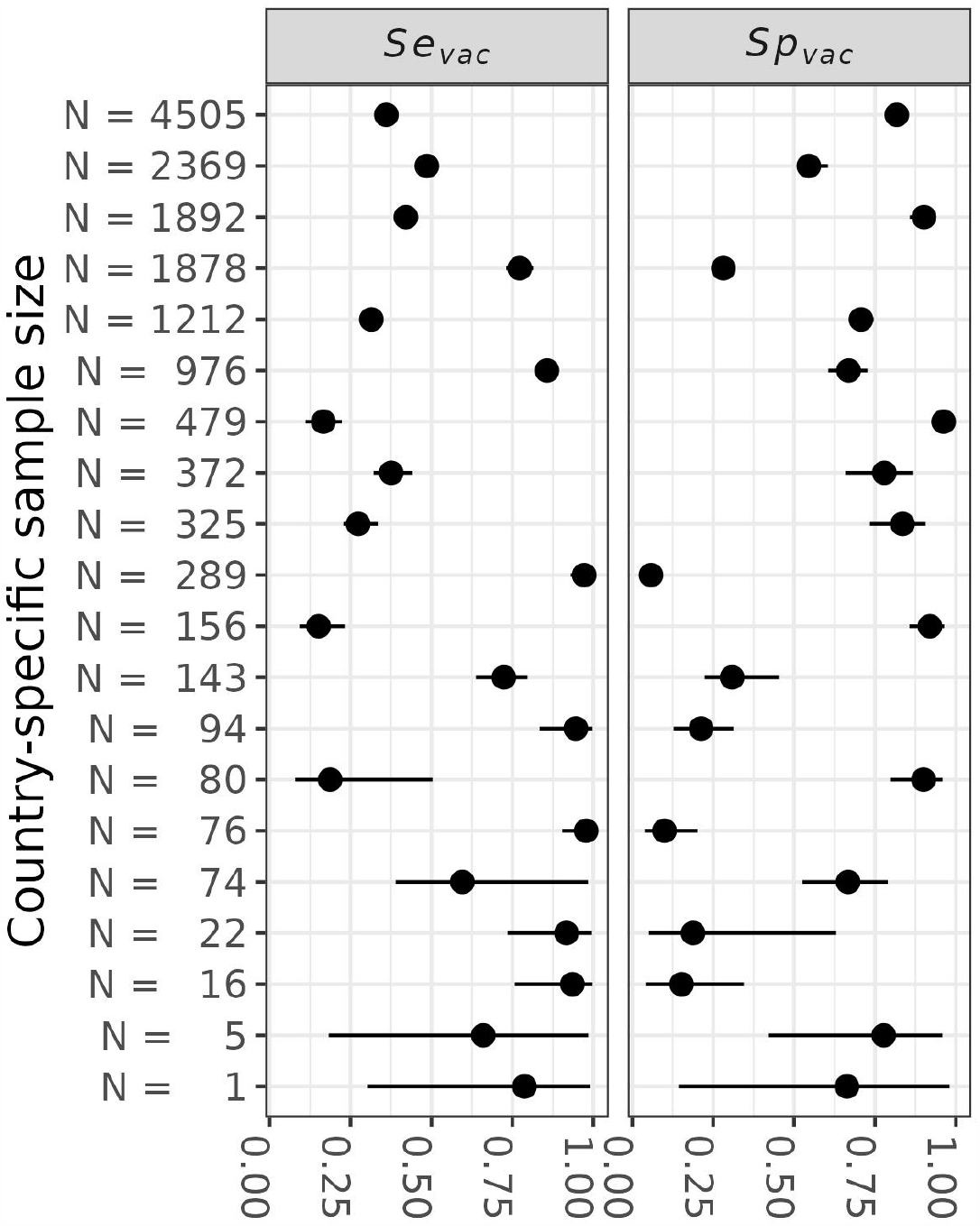
Relationship between the number of data points from each country (y-axis) and uncertainty in the estimates of *Se*_*vac*_ (left) and *Sp*_*vac*_ (right) for the default model. Points and lines represent the posterior medians and 95% credible intervals of *Se*_*vac*_ and *Sp*_*vac*_ for each country.

**Figure S2:**
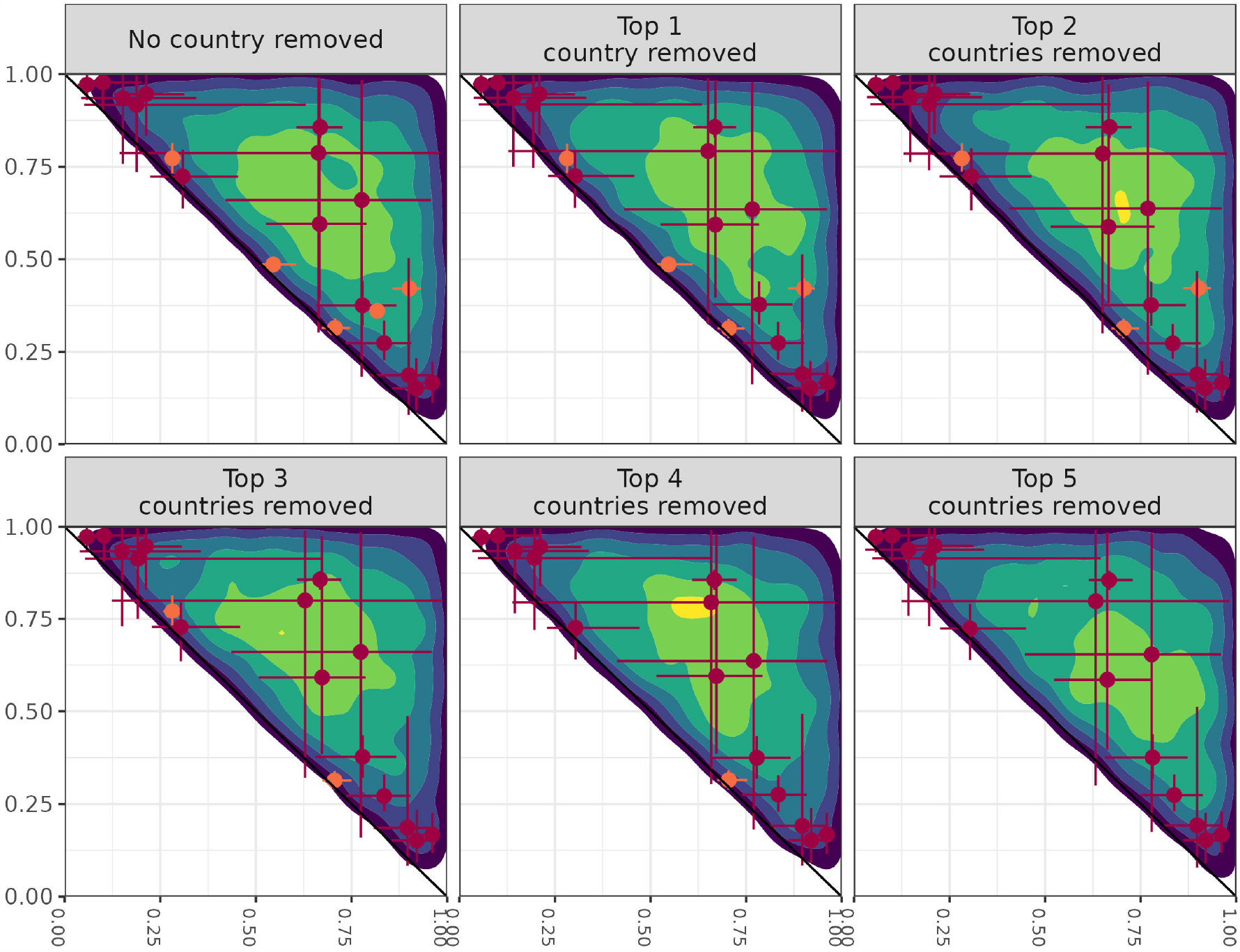
Estimates of *Se*_*vac*_ and *Sp*_*vac*_ from the default model after excluding the top five countries with the most data points. The dots and lines represent posterior medians and 95% credible intervals of country-specific estimates of *Se*_*vac,k*_ and *Sp*_*vac,k*_. The orange dots and lines indicate the top five countries with the most data points. Lighter shading indicates higher probability densities.

**Figure S3:**
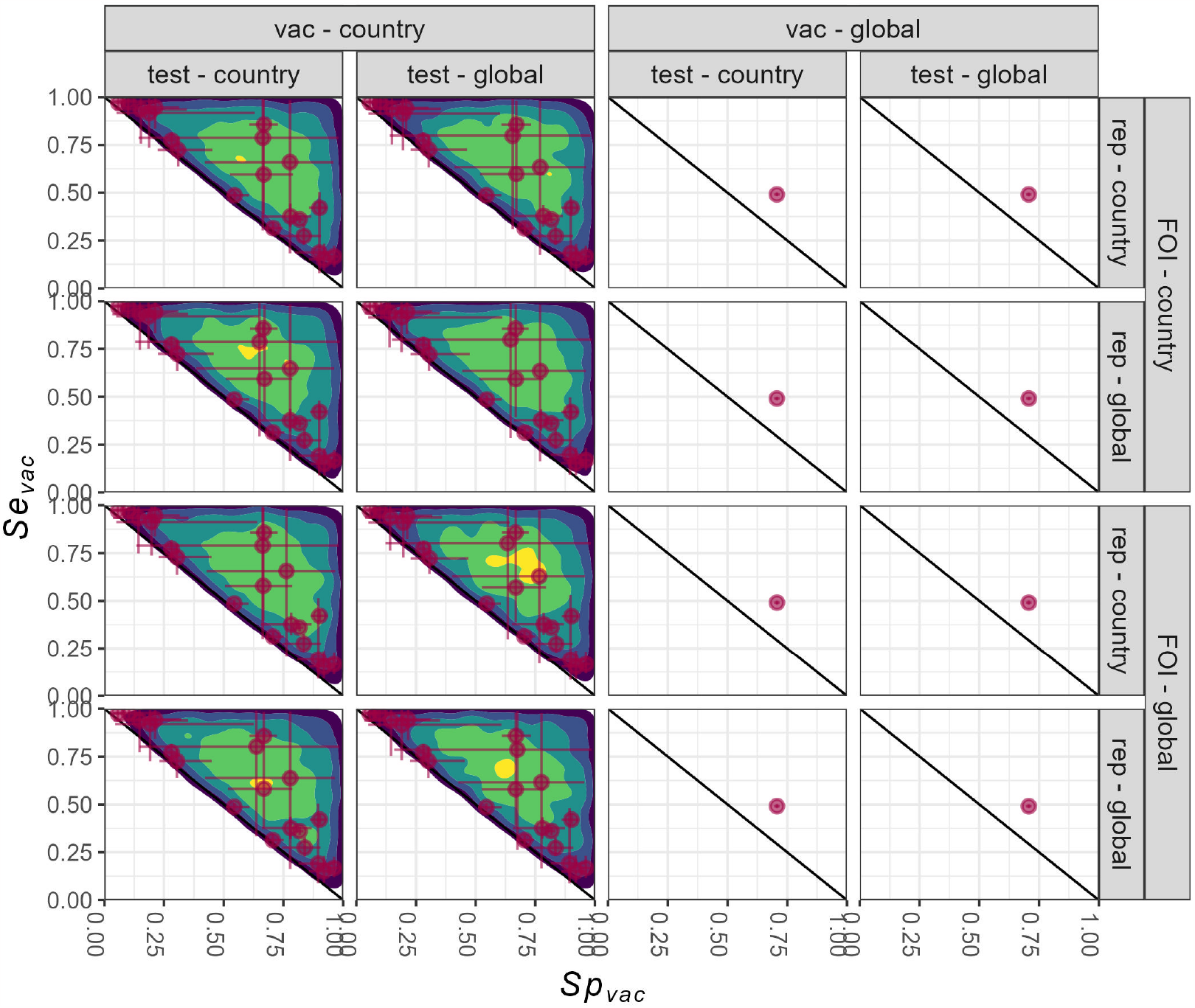
Estimates of *Se*_*vac*_ (y-axis) and *Sp*_*vac*_ (x-axis) from the 16 models (panels). In the columns and rows, geographic variability is indicated by whether parameters vary by country or do not vary and are thus global constants. The default model (top left) includes geographic variability in *Se*_*vac*_ and *Sp*_*vac*_ (vac), *Se*_*test*_ and *Sp*_*test*_ (test), force of infection (FOI), and reporting proportion (rep). The dots and lines represent posterior medians and 95% credible-intervals of country-specific estimates of *Se*_*vac,k*_ and *Sp*_*vac,k*_. Lighter shading indicates higher probability densities.

**Figure S4:**
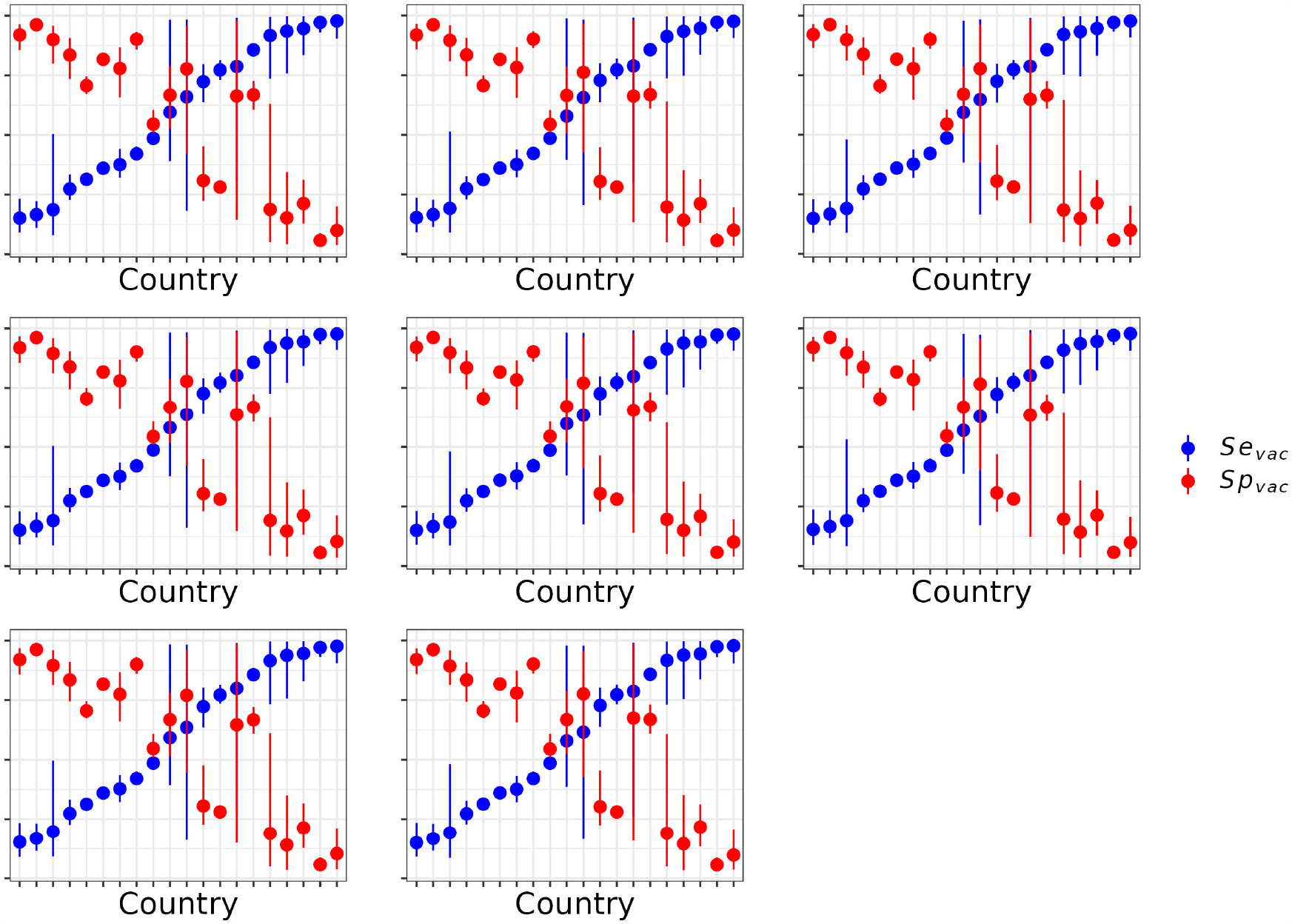
Marginal posterior distributions of *Se*_*vac*_ (blue) and *Sp*_*vac*_ (red) from the eight models (panels) that accounted for geographic variability in those parameters. The dots and lines represent medians and 95% credible intervals of country-specific estimates of *Se*_*vac,k*_ and *Sp*_*vac,k*_. The country-specific estimates are sorted in ascending order based on *Se*_*vac,k*_. A noticeable trend of negative correlation between *Se*_*vac,k*_ and *Sp*_*vac,k*_ is observed for all models.

**Figure S5:**
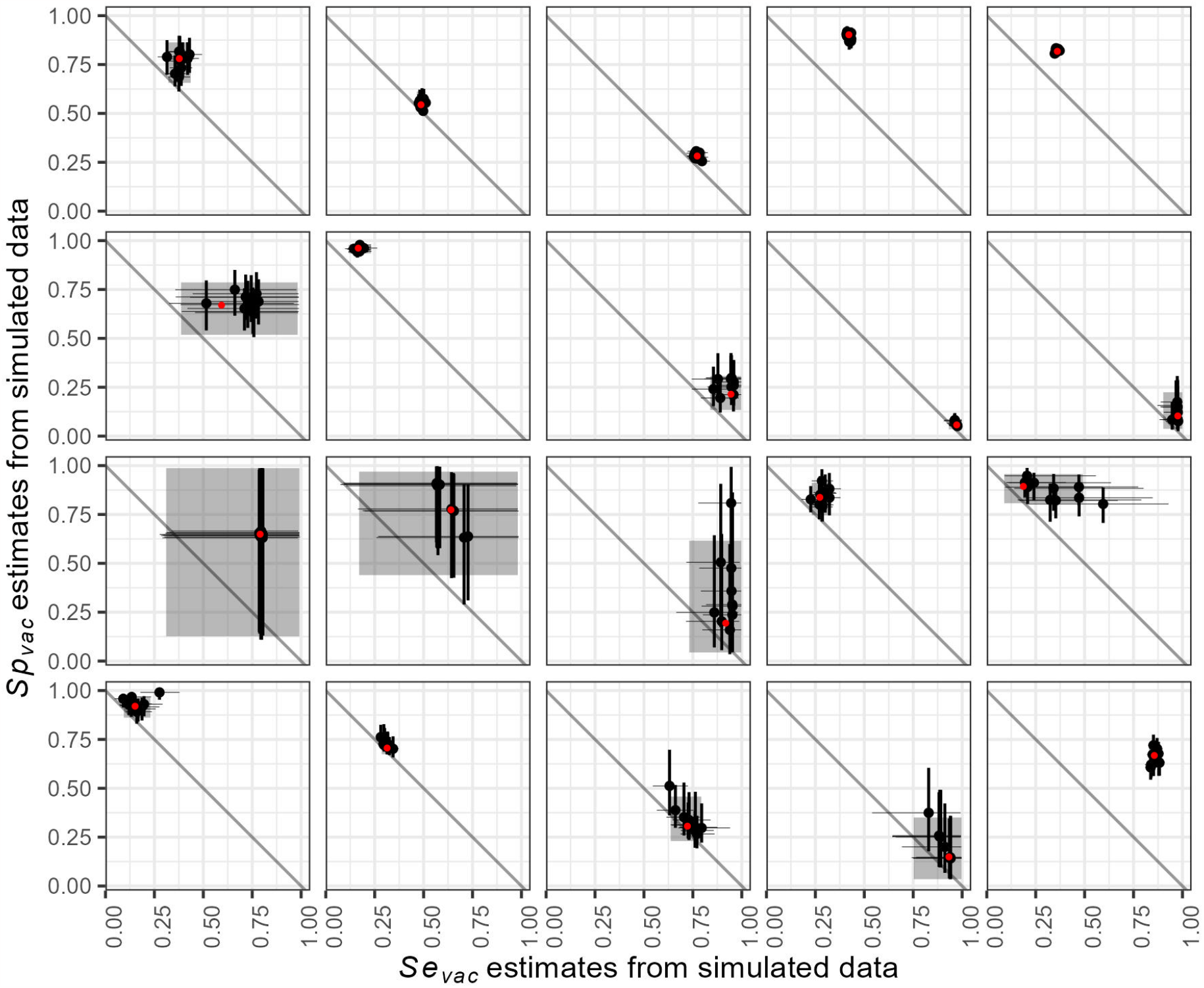
Estimation of *Se*_*vac,k*_ (x-axis) and *Sp*_*vac,k*_ (y-axis) for each of 20 countries (panels) from ten data sets simulated with posterior median parameter values. The ten black dots in each panel and their lines represent median estimates and 95% credible intervals obtained from our analysis of the ten simulated data sets. The red dots and gray areas represent median posterior estimates and their corresponding 95% credible intervals obtained from our analysis of empirical data. The red dots were also the parameter values used to generate the ten simulated data sets.

**Figure S6:**
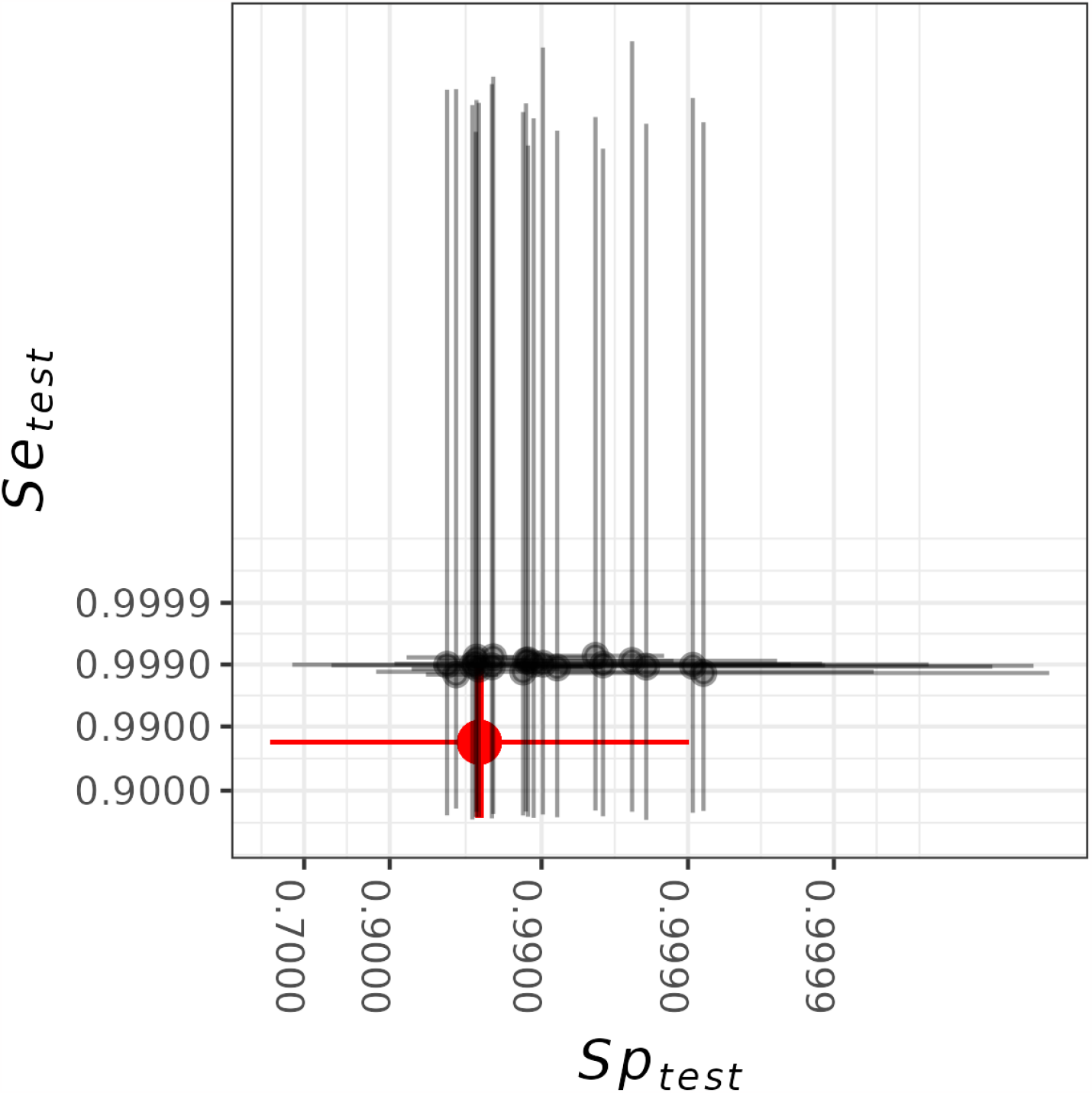
Estimates of *Se*_*test*_ and *Sp*_*test*_ from the default model. The points with their lines represent the median estimates and the 95% CrI of country-specific estimates *Se*_*test,k*_ and *Sp*_*test,k*_. The red points and lines indicate the medians and 95% uncertainty intervals of the priors used.

**Figure S7:**
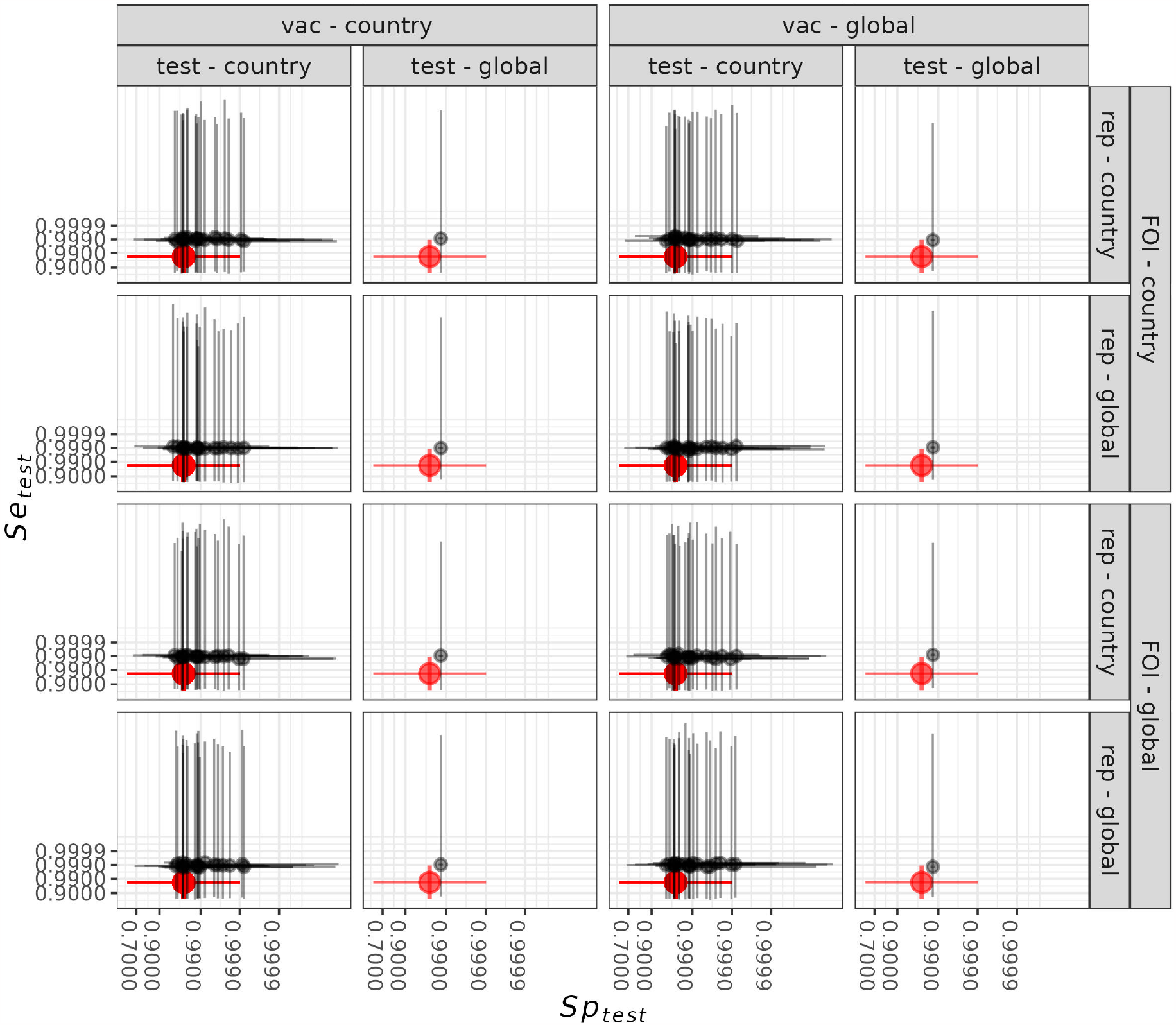
Estimates of *Se*_*test*_ (y-axis) and *Sp*_*test*_ (x-axis) from the 16 models (panels). In the columns and rows, geographic variability is indicated by whether parameters vary by country or do not vary and are thus global constants. The default model (top left) includes geographic variability in *Se*_*vac*_ and *Sp*_*vac*_ (vac), *Se*_*test*_ and *Sp*_*test*_ (test), force of infection (FOI), and reporting proportion (rep). The dots and lines represent posterior median estimates and the 95% credible intervals of country-specific estimates *Se*_*test,k*_ and *Sp*_*test,k*_. The red points and lines indicate the medians and 95% uncertainty intervals of the priors.

**Figure S8:**
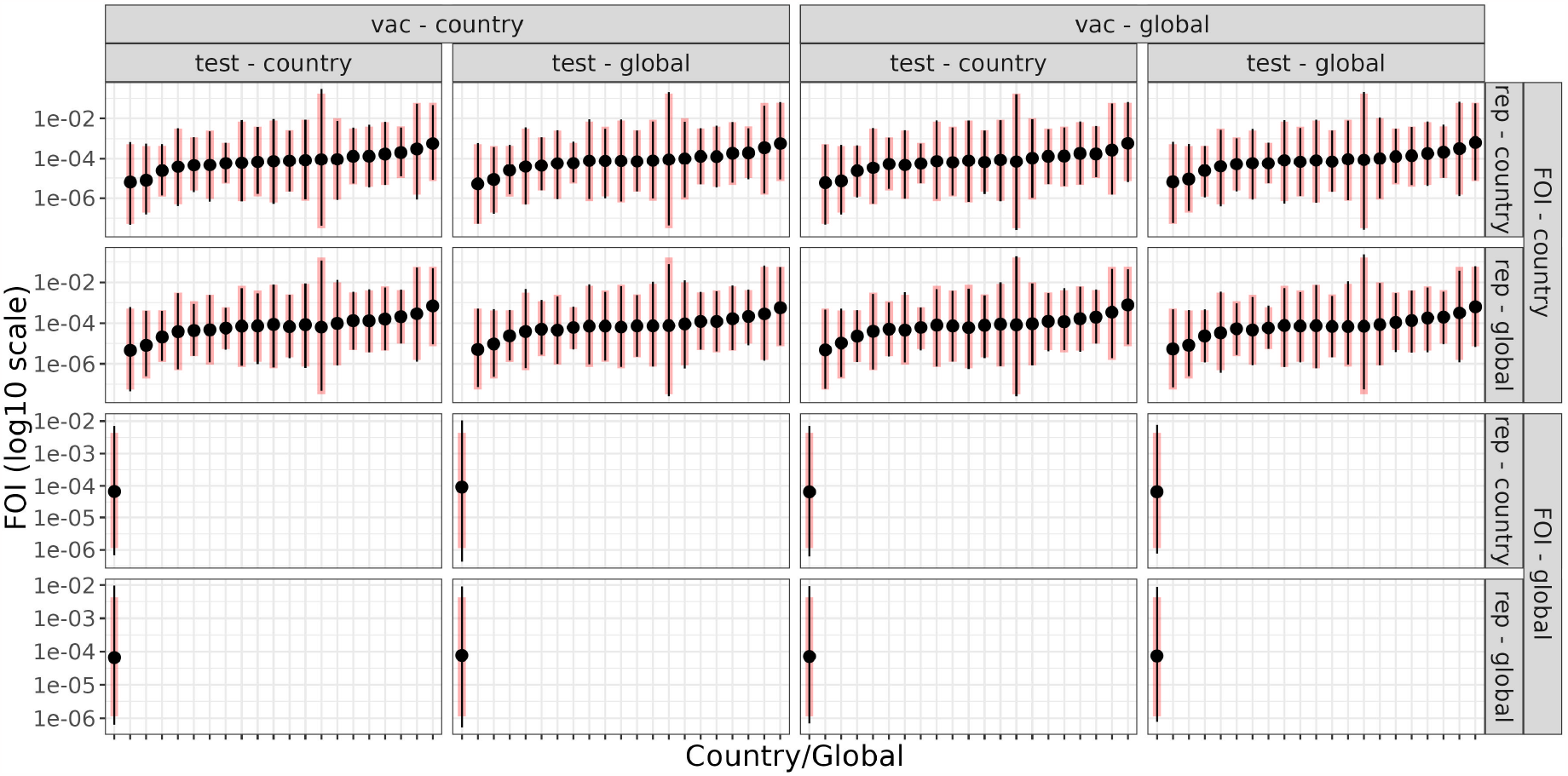
Estimates of force of infection, *λ*_*k*_, (y-axis) from the 16 models (panels). Dots and lines represent posterior medians and 95% credible intervals of *λ*_*k*_ for each country. Vertical red lines represent the 95% intervals of the prior distributions, which were obtained from a previous study [26]. The default model (top left) includes geographic variability in *Se*_*vac*_ and *Sp*_*vac*_ (vac), *Se*_*test*_ and *Sp*_*test*_ (test), force of infection (FOI), and reporting proportion (rep). In the columns and rows, geographic variability is indicated by whether parameters vary by country or do not vary and are thus global constants.

**Figure S9:**
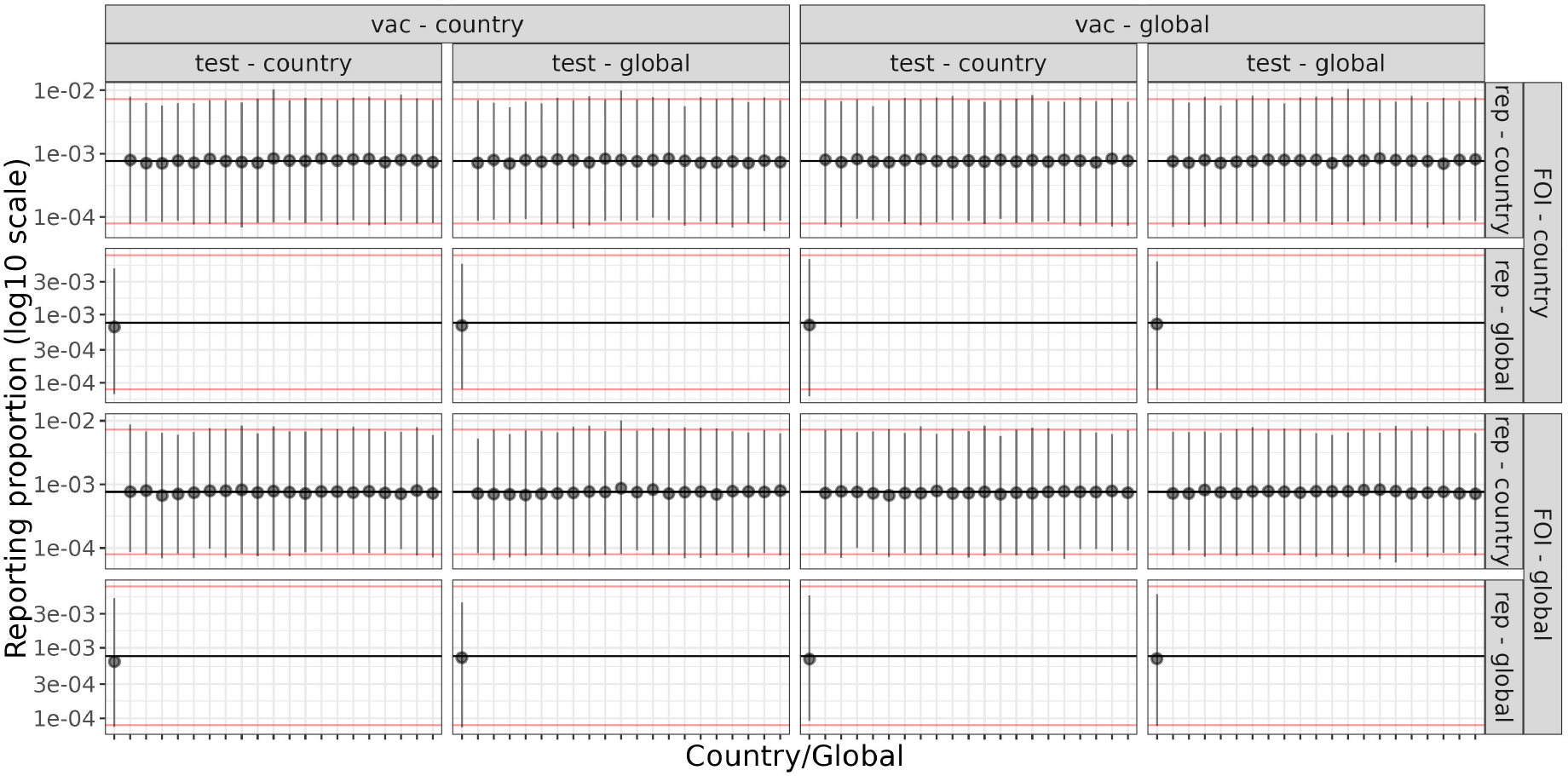
Estimates of reporting proportion, *ρ*_*k*_, from the 16 models (panels). Black dots and lines represent posterior medians and 95% credible intervals of *ρ*_*k*_ for each country (or for the global scale, depending on the model). Red lines represent the 95% interval and the horizontal black lines represent the median of the prior distribution, which was obtained from a previous study [26]. The default model (top left) includes geographic variability in *Se*_*vac*_ and *Sp*_*vac*_ (vac), *Se*_*test*_ and *Sp*_*test*_ (test), force of infection (FOI), and reporting proportion (rep). In the columns and rows, geographic variability is indicated by whether parameters vary by country or do not vary and are thus global constants.

**Figure S10:**
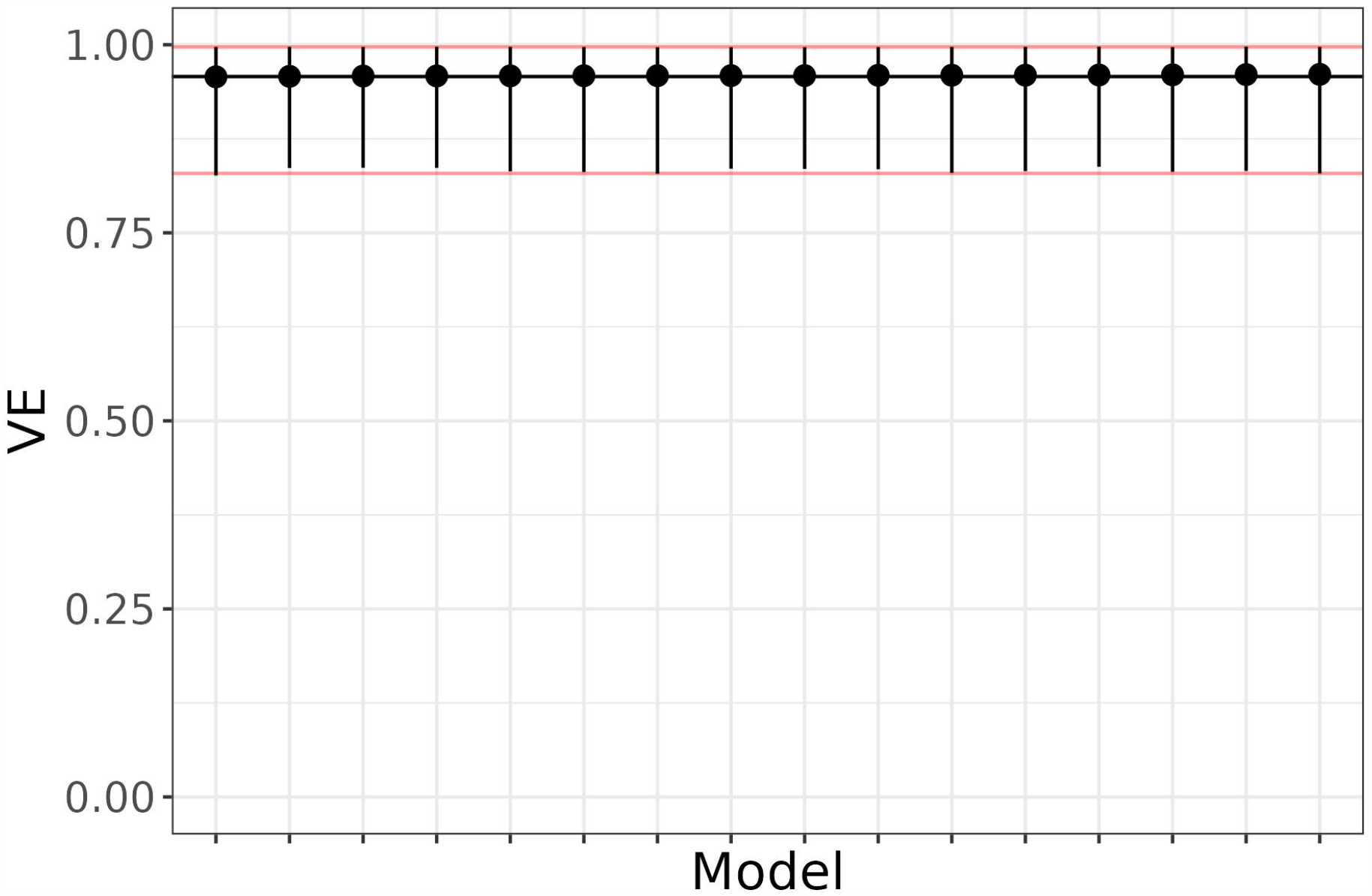
Estimates of vaccine efficacy, *V E*, (y-axis) from all 16 models (x-axis). Black dots and lines depict posterior medians and 95% credible intervals for *V E*. The horizontal red lines represent the 95% interval of the prior distribution, and the black line represents the median of the prior distribution.

**Figure S11:**
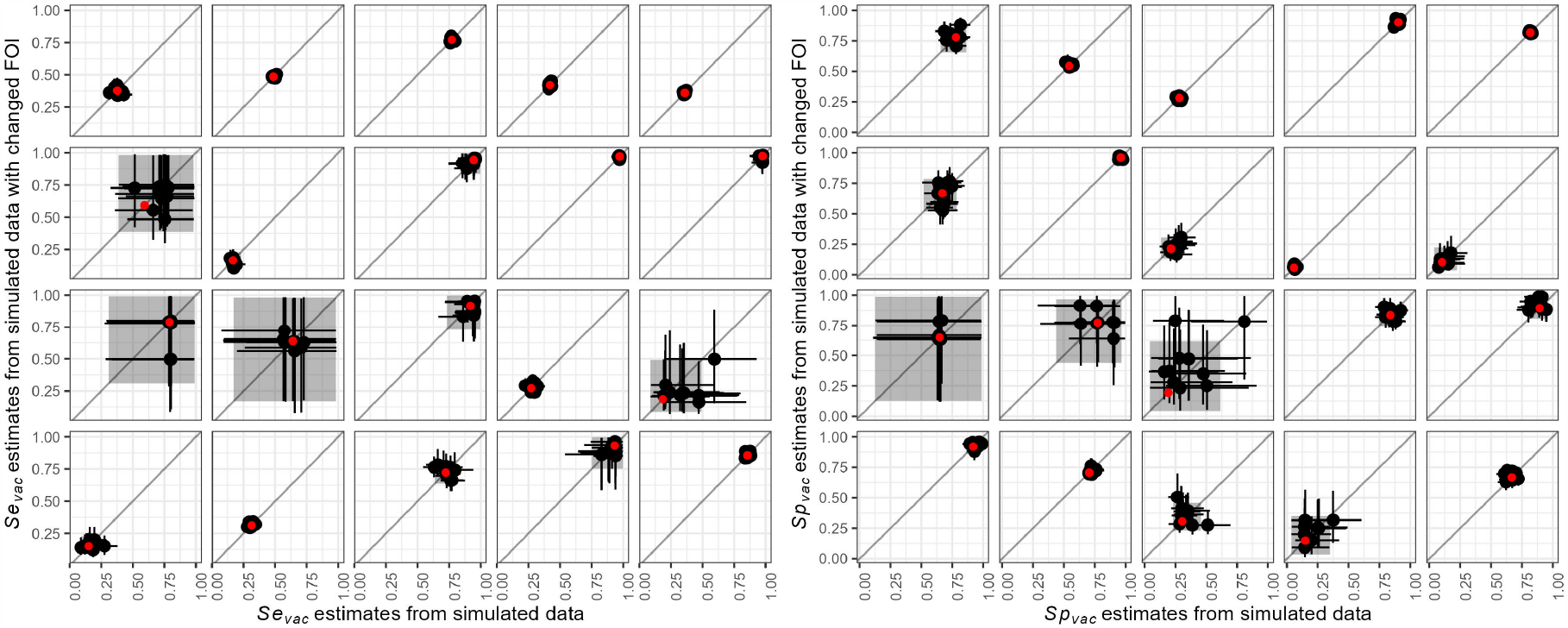
Estimates of *Se*_*vac,k*_ (left) and *Sp*_*vac,k*_ (right) from simulated data with (y-axis) or without (x-axis) changed values of *λ*_*k*_ for each of 20 countries (panels). In the unchanged scenario, we simulated ten data sets with posterior median parameter values. In the changed scenario, we simulated ten data sets with *λ*_*k*_ set to values of either 10^−*5*^, 10^−*4*^, 10^−*3*^, 10^−*2*^, or 10^−*1*^. The ten black dots in each panel and their lines represent median estimates and 95% credible intervals obtained from our analysis of the ten simulated data sets. The red dots and gray areas represent median posterior estimates and their corresponding 95% credible intervals obtained from our analysis of empirical data.

**Figure S12:**
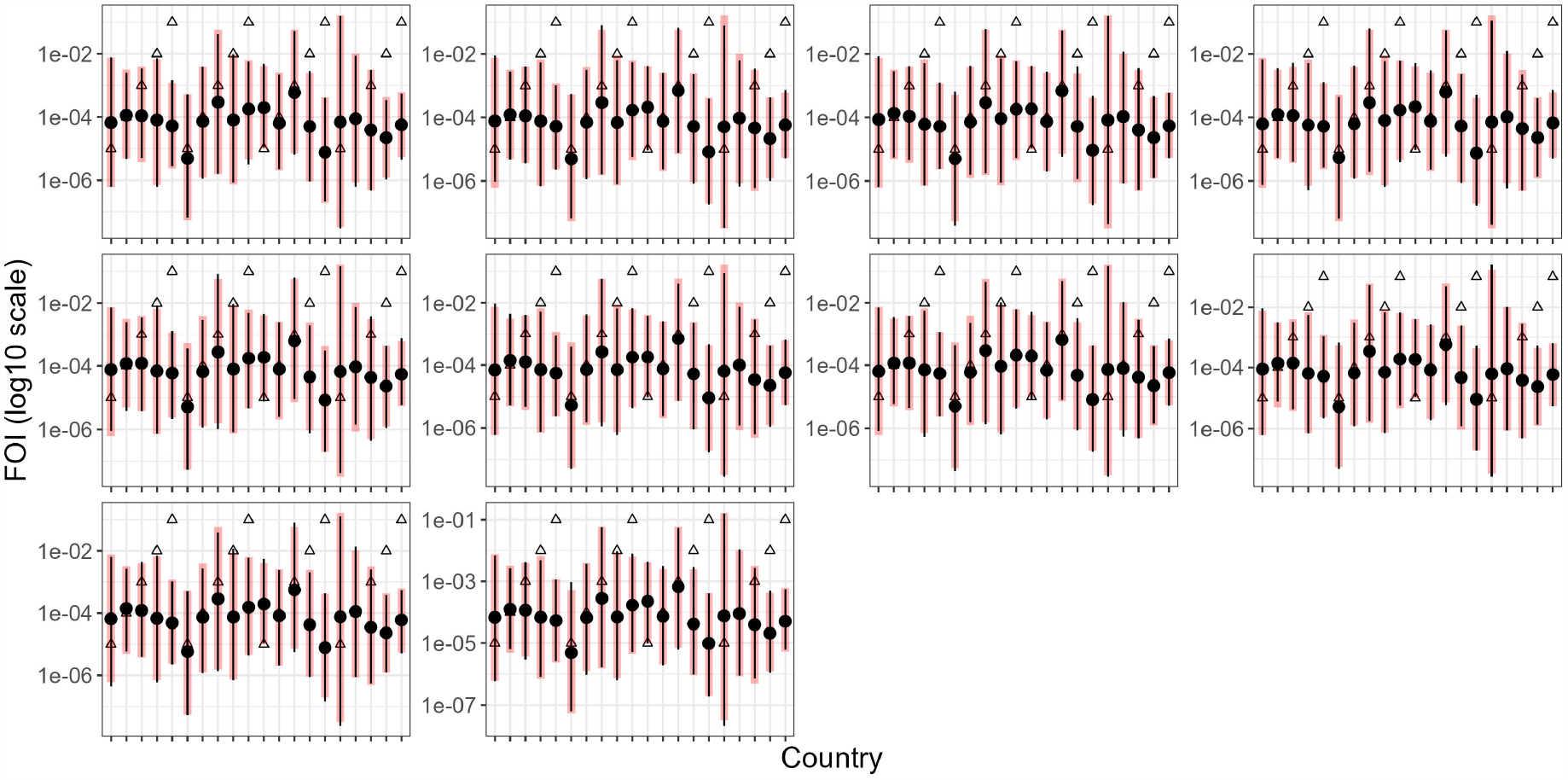
Estimates of force of infection, *λ*_*k*_, (y-axis) for each of 20 countries (x-axis) in ten simulated data sets (panels). Black dots and vertical lines denote posterior median and 95% credible intervals. Vertical red lines represent the 95% intervals of the prior distributions, sourced from a previous study [26]. Triangles correspond to the values of *λ*_*k*_ used to generate the simulated data.

**Figure S13:**
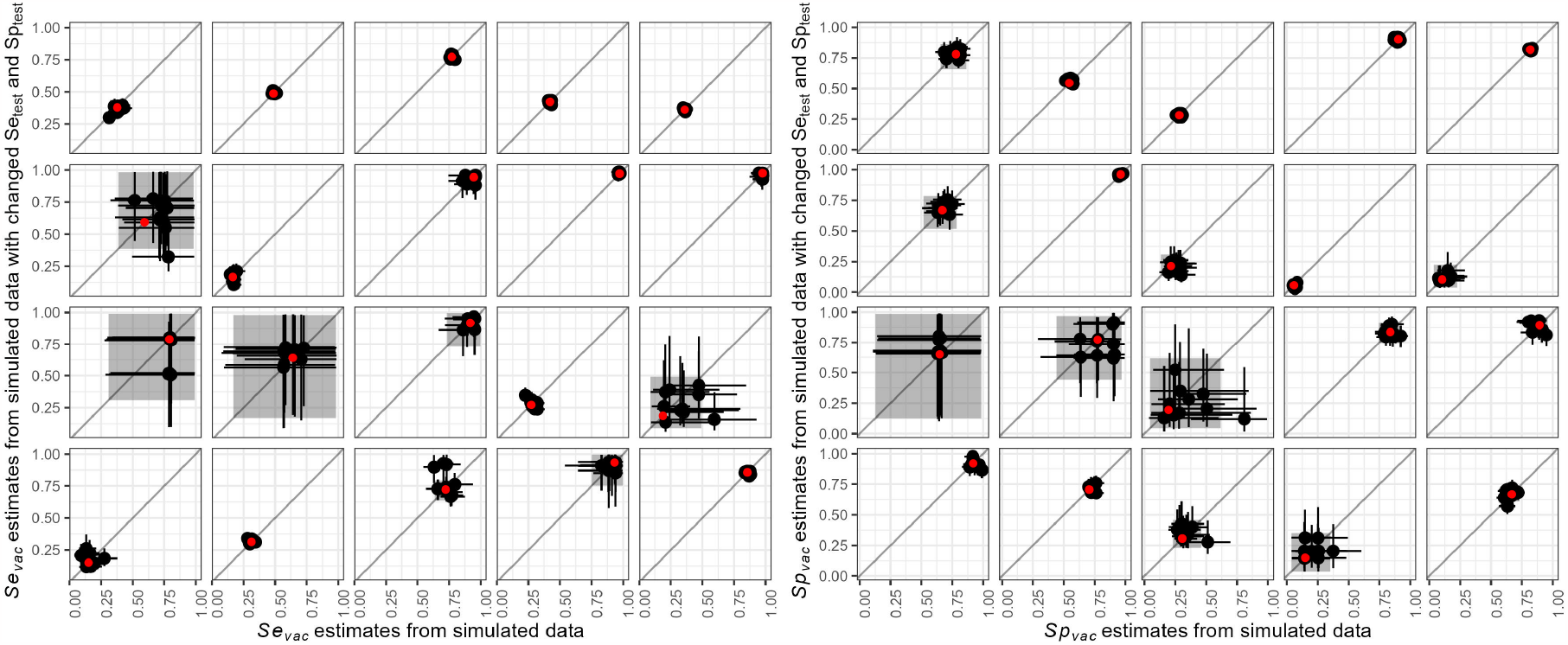
Estimates of *Se*_*vac,k*_ (left) and *Sp*_*vac,k*_ (right) from simulated data with (y-axis) or without (x-axis) changed values of *Se*_*test,k*_ and *Sp*_*vac,k*_ for each of 20 countries (panels). In the unchanged scenario, we simulated ten data sets with posterior median parameter values. In the changed scenario, we simulated ten data sets with *Se*_*test,k*_ and *Sp*_*test,k*_ set to 0.7. The ten black dots in each panel and their lines represent median estimates and 95% credible intervals obtained from our analysis of the ten simulated data sets. The red dots and gray areas represent median posterior estimates and their corresponding 95% credible intervals obtained from our analysis of empirical data.

**Table S1:**
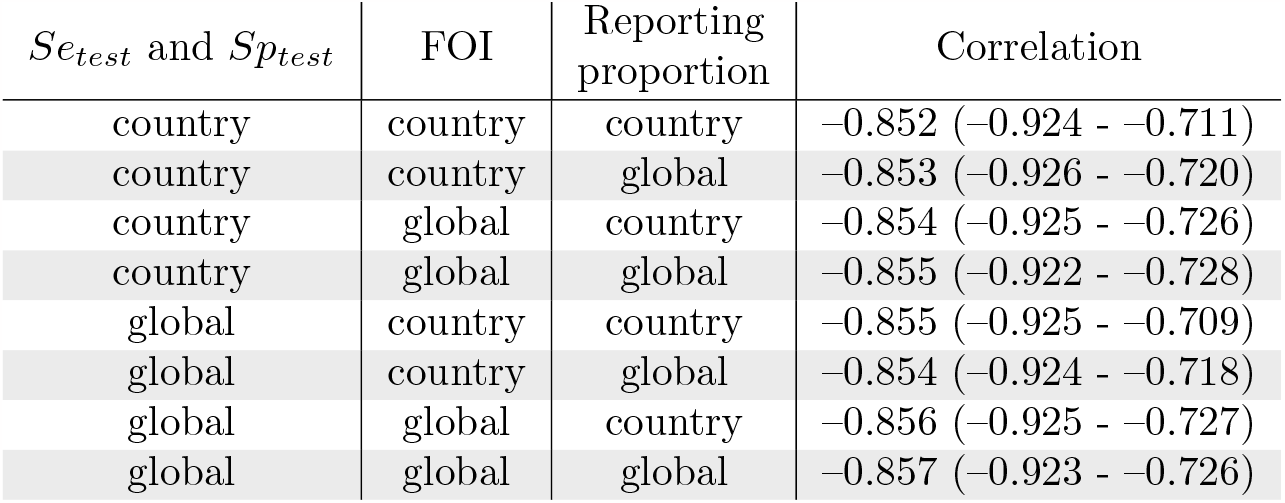
Correlation between *Se*_*vac,k*_ and *Sp*_*vac,k*_ across countries in eight models that account for geographic variability in *Se*_*vac*_ and *Sp*_*vac*_.

